# Accuracy of Samsung Smartphone Integrated Pulse Oximetry Meets Full FDA Clearance Standards for Clinical Use

**DOI:** 10.1101/2021.02.17.21249755

**Authors:** Sara H. Browne, Mike Bernstein, Philip E. Bickler

## Abstract

**Background:** Pulse oximetry is used as an assessment tool to gauge the severity of COVID-19 infection and identify patients at risk of poor outcomes. ^1,2,3,4^ The pandemic highlights the need for accurate pulse oximetry, particularly at home, as infection rates increase in multiple global regions including the UK, USA and South Africa ^5^. Over 100 million Samsung smartphones containing dedicated biosensors (Maxim Integrated Inc, San Jose, CA) and preloaded Apps to perform pulse oximetry, are in use globally. We performed detailed in human hypoxia testing on the Samsung S9 smartphone to determine if this integrated hardware meets full FDA/ISO requirements for clinical pulse oximetry.

**Methods:** The accuracy of integrated pulse oximetry in the Samsung 9 smartphone during stable arterial oxygen saturations (SaO_2_) between 70% and 100% was evaluated in 12 healthy subjects. Inspired oxygen, nitrogen, and carbon dioxide partial pressures were monitored and adjusted via a partial rebreathing circuit to achieve stable target SaO_2_ plateaus between 70% and 100%. Arterial blood samples were taken at each plateau and saturation measured on each blood sample using ABL-90FLEX blood gas analyzer. Bias, calculated from smartphone readings minus the corresponding arterial blood sample, was reported as root mean square deviation (RMSD).

**Findings:** The RMSD of the over 257 data points based on blood sample analysis obtained from 12 human volunteers tested was 2.6%.

**Interpretation:** Evaluation of the smartphone pulse oximeter performance is within requirements of <3.5% RMSD blood oxygen saturation (SpO_2_) value for FDA/ISO clearance for clinical pulse oximetry. This is the first report of smartphone derived pulse oximetry measurements that meet full FDA/ISO accuracy certification requirements. Both Samsung S9 and S10 contain the same integrated pulse oximeter, thus over 100 million smartphones in current global circulation could be used to obtain clinically accurate spot SpO_2_ measurements to support at home assessment of COVID-19 patients.

## Introduction

Pulse oximetry supports the triage and initial management of symptomatic adults during respiratory infection pandemics^6,7^. Pulse oximetry is being used during the current pandemic as an assessment tool to identify patients with COVID-19 at risk of poor outcomes ^1^ and gauge the severity of infection.^2,3,4^ The pandemic has highlighted the need for accurate pulse oximetry, particularly at home. In conjunction with COVID symptom diaries, at home oximetry is being used to detect early deterioration of patients in primary and community care settings.^8,9^ Primary care assessment pathways within major healthcare systems have developed guidance directives centered around SpO_2_ measurements taken at home due to isolation; these readings with respiratory rates (RR) and mental state assessments obtained during virtual visits are being used to generate national early warning scores (NEWS).^8^

Remote healthcare providers also guide patients on early warning signs of worsening COVID-19 infection centered on deteriorating at home SpO_2_ measurements over time, as well as exertion oximetry (exercise induced hypoxia). ^8,10,11^ The term ‘silent hypoxia’ associated with COVID-19 infection has appeared in the literature to describe the lack of dyspnea in some patients. ^10^ These reports serve as a reminder that the diagnosis of hypoxemia, even severe hypoxemia, is sometimes missed without objective measures such as non-invasive oximetry or arterial blood gases (ABGs).^11^

The broad availability of accurate pulse oximetry is evidently of considerable significance during the current pandemic; furthermore, inequity in the distribution of accurate oximetry devices globally has been well documented by the WHO^12^. FDA/ISO cleared pulse oximeter devices are expensive and it is likely that the majority of at home pulse oximeters currently in use, including those dispensed by large healthcare systems, are inexpensive devices that may be associated with wide variability in accuracy.^13^

Samsung Galaxy S9 and 10 smartphones include the same dedicated hardware using high grade integrated biosensors and preloaded proprietary Samsung Apps that process the sensor data, calculate saturation and heartrate and then display the user’s pulse oximetry measurement ^14^. In 2018, an estimated 45 million S9 smartphones were shipped worldwide. ^15^ In addition reports indicate that 71 million S10 smartphones were shipped by the end of 2019. Given the increased global demand for accurate pulse oximetry associated with the COVID-19 pandemic, we performed detailed in human testing to determine if these smartphones meet the FDA/ISO requirements for clinical pulse oximetry. FDA 510K clearance and ISO CE marking require assessment of accuracy during hypoxemia. This type of testing was introduced by John Severinghaus MD of the University of California, San Francisco (UCSF) and refined by Dr. Philip Bicker UCSF^16,17^ During this testing detailed evaluation of bias between smartphone pulse oximeter readings and arterial blood gas measurements are made.

## Methods

A Samsung S9+ smartphone (Samsung, Seoul, South Korea) containing Maxim Integrated biosensors, part number MAX86916 (Maxim Integrated, San Jose, CA) associated with the proprietary Samsung Health App was the test instrument. The test instrument has phone biosensors include a specialized dedicated photodetector and 2 precision wavelength LEDs equivalent to those used in clinical oximeters that in this case are dedicated to the oximetry function (See Figure 1). The associated App processes the sensor data, calculates saturation and heartrate then displays it for the user view and record if desired.

**Figure 1:**
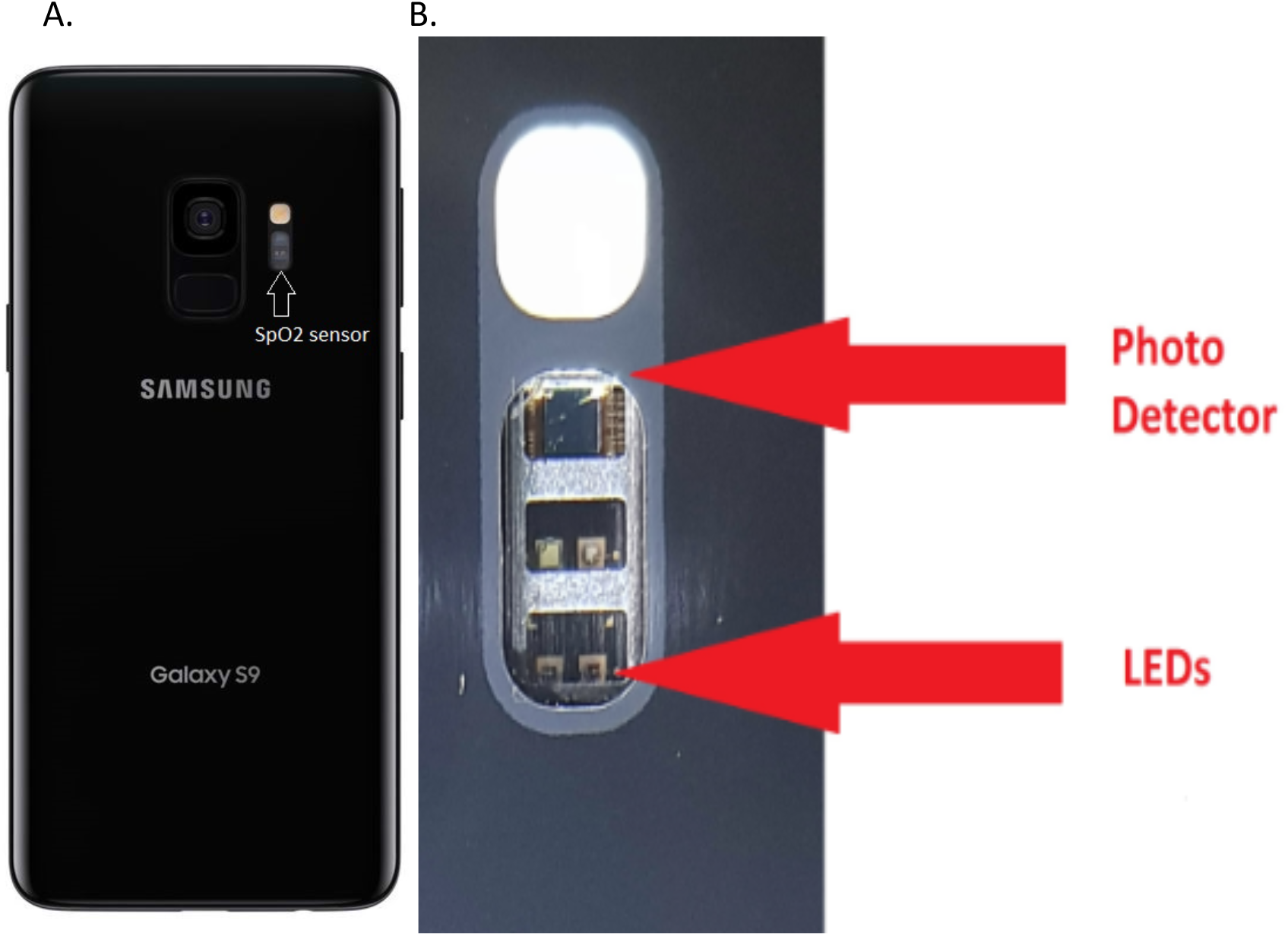
(A) shows the location of the SpO_2_ sensor on the back of the smartphone making clear that the sensor is entirely separate from the other phone devices such as the camera. In the expanded view (B) the dedicated photo sensor and LEDs for the pulse oximetry function are shown. The device has a total of 4 LEDs two of which are reserved for future additional functionality. The arrow indicates the 660 nM red and 910nM infrared LEDs used in this measurement. The photodetector is connected to an extremely low noise analog channel allowing measurement on a broad range of skin colors.

Evaluation of test instrument oximeter performance was done using controlled steady-state hypoxia at the UCSF Hypoxia Research Laboratory. The work was sanctioned by the UCSF Committee on Human Research, protocol 10-00437, and conformed to all internationally accepted standards for the protection of human subjects. We enrolled 12 participants, 4 male, 8 female, including 3 participants with darkly pigmented skin. All volunteers provided written informed consent. The left forefinger of each subject was placed over the smartphone sensor system (see Figure 1). During laboratory testing a series of desaturations are performed over 30 minutes. To enable test Participant to hold their finger continuously in position over a 30 minute period of time a silicone boot was used during Laboratory testing. Figure 2 shows the silicone boot attached to a plastic cell phone case that was utilized in this test.

**Figure 2:**
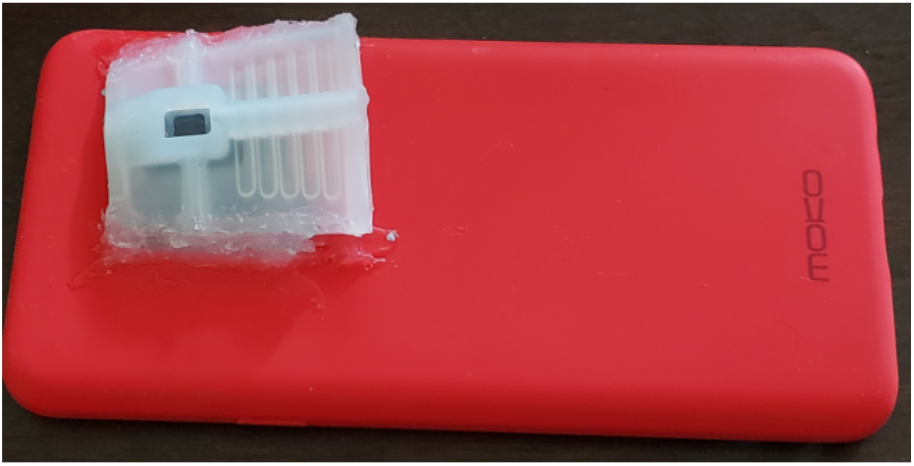
Plastic phone case with silicon boot attached used to hold each participant’s finger in position continuously for over 30 minutes.

A 22-gauge radial artery cannula was placed in each participant after lidocaine local anesthesia. The participants then breathed a mixture of air, nitrogen and carbon dioxide to produce stable levels of arterial saturation between 100% and 70%. When oxygenation was stable, as assessed from end-tidal gas and reference pulse oximeters, 2 arterial blood samples were taken, 30 seconds apart. Saturation and other oximetry parameters was measured on each blood sample with an ABL-90FLEX blood gas analyzer (Radiometer, Brea, CA, USA).

Statistical Analysis: Bias was computed as smartphone readings minus the corresponding arterial blood sample value. Bias is reported as root mean square deviation (RMSD). A plot was generated following Bland and Altman with adjustments for multiple measurements for each individual according to the “Method Where the True Value Varies^.”17^ The FDA 2013 guidance prescribes the use of a Bland Altman error plot with specifically calculated limits of agreement.^18^

## Results

In this study no plateaus were rejected for lack of stability between the first blood sample and the second. Eighteen readings from the device under test were rejected because the results were delayed more than 15 seconds past the sample time. Twelve subjects completed the study and there were no adverse events.

Figure 3 displays a modified Bland-Altman Plot of the entire data set. The RMSD of the over 257 data points based on blood sample analysis obtained from 12 human volunteers tested is 2.6%.

**Figure 3:**
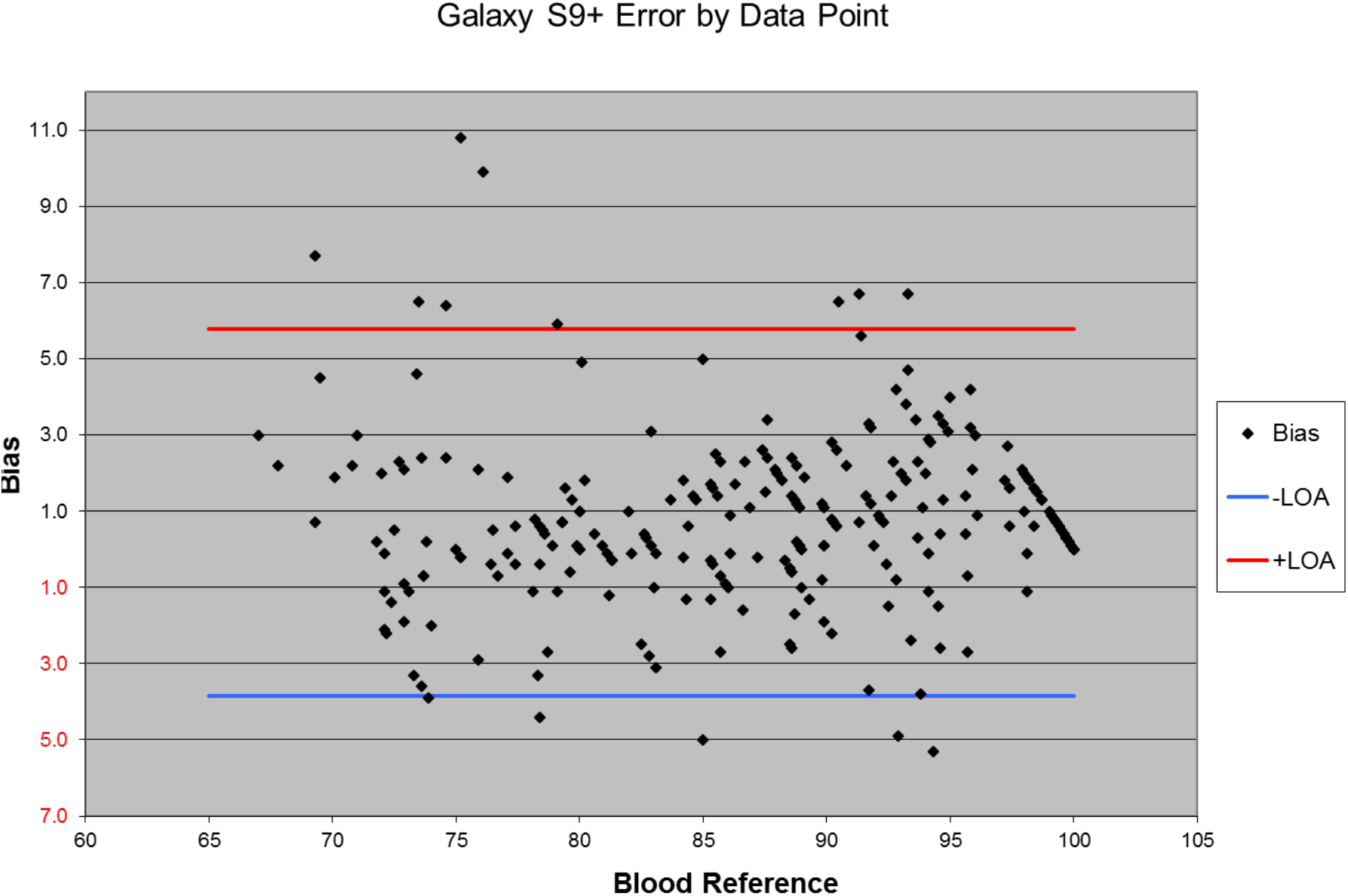
Plot of data collected from 12 human volunteers during this test of the bias in blood oxygen measurements. Each point on the plot represents paired blood oxygen levels recorded by the hemoximeter associated with the Samsung Phone reading. The upper and lower limits of agreement per Bland Altman 2007^17^ are shown in red and blue lines respectively.

## Discussion

### Interpretation of findings

Evaluation of oximeter performance during controlled steady-state hypoxia using the Samsung S9+ smartphone, containing Maxim Integrated biosensors and Samsung Health App, revealed that readings had an RSMD of 2.6% from over 257 simultaneous ABGs. This finding is well within requirements for FDA/ISO clearance for clinical pulse oximetry, which is <3.5% RMSD SpO_2_ value. This is the first report of smartphone derived pulse oximetry measurements that meets full FDA/ISO accuracy certification requirements. These findings indicate that an estimated 45 million Samsung S9/9+ smartphones, with existing embedded dedicated hardware and preloaded Apps, in current circulation may be used to take accurate clinical grade pulse oximeter readings. Furthermore, S10 series smartphones contain the same biosensor hardware and preloaded Apps with the same proprietary Samsung algorithm, with reports stating 71 million such smartphones were shipped by the end of 2019. ^19^ Consequently, the implications of our study for global access to accurate pulse oximetry during the current COVID-19 pandemic are considerable.

Up to this point studies evaluating smartphone pulse oximetry have shown variable accuracy with the minority supporting any clinical use. Jordan et al. compared pulse oximetry Apps downloaded on to iPhones to reference monitors within an Emergency room setting and found the Apps provided inaccurate measurements with a sensitivity for detection SpO_2_<94% on the reference monitor ranging from 0-69%.^20^ Two of these Apps utilized the onboard light and camera lens (Pox and Ox) and one used an external device that plugged into the iPhone. Alexander et al., ^21^ evaluated two other smartphone Apps (Pulse Oximeter and Pulse Oximeter Pro) again downloaded onto an iPhone and using onboard light and camera lens to perform SpO_2_ measurements, predominantly in a Pre-operative setting. Comparison of these Apps with clinical devices utilized by the Dept of Anesthesia showed similar mean and median values, but wide and highly significant variance in measurements. ^21^ Tayfur et al., compared SpO_2_ measurements obtained on 101 hospitalized patients (43% having pulmonary disease) using Samsung Galaxy S8 with that measured by simultaneous ABGs and reported measurements as highly correlated and having a small bias with narrow levels of agreement. ^22^ Modi et al. conducted 6 minute walk pre- and post-oximetry testing to perform a comparison of a Massimo-radical7 device, a clip Kenek sensor connected to an iPhone, and a Samsung Galaxy 8. ^23^ They reported a correlation between SpO_2_ measurements (r= 0.62 - 0.72, p<0.001) and provided values from 28 out of 47 participants on a Bland Altman analysis that used an average of the SpO_2_ obtained by the FDA approved Massimo-radical7 reference device and the smartphone readings as ‘true’ SpO_2_, making this data difficult to evaluate. Most recently, Browne et al reported findings from clinical studies on 320 participants that utilized a repeated measure, nested factorial design to evaluate the accuracy and precision of a smartphone model, containing Maxim Integrated biosensors and App in comparison to Welch Allyn reference units.^24^ Their results indicated a wide range of adult persons utilized a smartphone model to perform repeated pulse oximeter spot checks with an accuracy and precision equivalent to hospital grade clip sensors. ^24^ This clinical study had few hypoxemic patients, but did report human steady state hypoxia testing of the smartphone model versus a portable reference device that indicated FDA/ISO requirements were likely to be met if full testing was conducted.^24^

These widely varying findings are associated with studies that did not follow uniform protocols, were in different patient populations and none performed as we did full FDA/ISO required human hypoxia testing using ABGs as the oxygen saturation reference. Aside from these limitations, the major determinant of accuracy differences observed across these studies likely reflects differences in the quality of the underlying hardware and proprietary algorithms within the smartphone tested. Whilst traditional pulse oximeters in hospital settings rely on the transmission of light through cutaneous tissues such as the finger or ear lobe, smartphones systems utilize reflected light detected by a sensor on the same surface as the emitter. ^25^ Within such systems how the reflected light signal is obtained and analyzed is critical. Poorly performing pulse oximetry Apps used the onboard light and camera lens to obtain reflected light to detect PPG signals and conformational changes associated with hemoglobin binding. Signal obtained from camera associated sensors have relatively high signal to noise ratios and may even block near infrared light. ^25^ In contrast, the smartphone evaluated in our study has dedicated hardware specifically for pulse oximetry function (see Figure 1) with 660nM red and 910nM infrared LEDs used to take measurements, a photodetector connected to an extremely low noise analog channel allowing measurement on a broad range of skin colors, and placement entirely separate from other phone devices such as the camera. Studies testing similar hardware and algorithms reported high SpO_2_ measurement accuracy. ^24, 22^ All future studies evaluating smartphone oximetry accuracy should specify the type of embedded hardware including biosensor manufacturer, App proprietor and site of placement within the smartphone, as these features constitute the actual oximetry device

### Implications for Clinical Assessment

Our introduction described the widespread health system guidance on the use of at home SpO_2_ measurement to grade the severity of COVID-19 infection. Currently, infection numbers are increasing rapidly in association with the presence of the UK variant in multiple regions globally, resulting in greater demand for effective, inexpensive at home monitoring to identify patient deterioration^26,27^. Media reports indicate consumer purchase of oximeters has increased considerably since the start of the current pandemic. ^25^ The foremost concern of experts regarding home pulse oximetry during the current pandemic is the accuracy of SpO_2_ measurements, particularly as saturation falls below 90%.^25^ Data on the accuracy of inexpensive stand-alone oximeters is limited, as there has been little regulatory oversight.^13,25^ The best study available on the accuracy of inexpensive finger oximeters reported wide variability, with the majority of devices tested demonstrating highly inaccurate readings during hypoxia.^13^ Consumers are essentially unaware of these differences and healthcare workers only find reliable accuracy information for more expensive devices (>$150) on the market. ^25^ As the current cost of FDA cleared pulse oximeters varies from hundreds to greater than a thousand USD, it is highly likely the majority of at home pulse oximeters in use are not FDA/ISO cleared. In this regard our findings indicate that pulse oximetry measurements obtained by the Samsung Galaxy S9/9+ meet prescribed standards for clinical use and are more accurate than stand-alone oximeters for which no reliable information is available.

Pulmonologist practical guidance during COVID-19 at home monitoring states oximetry devices should provide some indication of pulse signal strength and that measurements should be taken and recorded two to three times a day at rest. ^25^ Using any oximeter, the finger must be positioned accurately relative to the sensor. The Samsung Healthcare App does provide assessment of adequate finger placement and adequate pulse strength and does not take a reading unless both are appropriate. This feature is important as accurately positioning the finger over the sensor on the back of the phone, then turning the hand to view the reading on the front, is more difficult than using a finger clip sensor. The oximetry function on Samsung S9/9+ smartphones is designed for spot check only, where the user holds their finger in place for approximately 30 seconds and gets a single reading. While multiple spot checks during the day can be made, the manner in which these readings are displayed under a label of ‘stress test’ within the Samsung Health App may be confusing to patients. However, the App does contain a ‘share’ function which enables approved providers to receive daily SpO_2_ and HR data that can be used to support at home assessment; and historical data is stored. Collaboration between software developers and smartphone manufacturers could easily improve this design to include instruction to take 2-3 readings a day at rest and warm extremities before measurement, in line with recommendations^25^, as well as present graphical data trends over time for transmission to healthcare practitioners.

### Study Limitations

This study used healthy volunteers under steady state hypoxia testing (SpO_2_ approximately 70-100%) and thus provides no data on accuracy below 70%. Data from healthy volunteers provides no information on accuracy of SpO_2_ measurement under conditions of reduced extremity pulsatile blood flow, associated with vasoconstriction due to Raynauds, hypotension or peripheral vascular disease, the latter is common in persons with diabetes and coronary heart disease, both risk factors for severe Covid-19 infection ^28^. Furthermore, measurement accuracy in the presence of diseases associated with hemoglobinopathies, such as sickle cell or thalassemia, or where elevated carboxyhemoglobin (potentially present in heavy smokers) or methemoglobin (associated with use of chloroquine, sulfonamides) my occur, is unknown. This study does not provide any evidence on the use of this system in children. Finally, the system tested is designed to provide SpO_2_ spot checks only, there is no evidence from our study to support use for continuous pulse oximetry.

## Conclusion

Based on our findings and the substantial need for widespread global access to accurate pulse oximetry during the current pandemic we recommend the high-grade biosensors and Samsung proprietary algorithm embedded in S9 and S10 smartphones pursue FDA 510K clearance and ISO certification for use to obtain spot SpO_2_ measurements for clinical use. Review could be prioritized and fast tracked by the FDA/ ISO organizations under the ‘Emergency Use Authorizations’ for ‘Remote or Wearable Patient Monitoring Devices associated with the Covid-19 pandemic’. ^30^ This may substantially support global efforts to respond to the current Covid-19, particularly in LMIC settings, where access to accurate spot pulse oximetry is limited ^11^.

## Data Availability

Full access to both the reference hemoximeter readings and paired readings from the test device can be obtained by emailing requests to

## Acknowledgements

Funding was provided by NIH grant supplement R01MH110057-04S to SHB and by the non-profit, Specialists in Global Health (https://sigh.global/). We thank Samsung Electronics, Suwon-si, Korea, for critical appraisal of this manuscript.

FIGURES: SMARTPHONE INTEGRATED PULSE OXIMETER

## Notes

### Competing Interest Statement

Sara Browne - no competing interests
Phil Bickler - no competing interets
Mike Bernstein, PhysioMonitor - Physiomonitor specializes in the testing and evaluation of pulse oximeters. In this research PhysioMonitor provided testing expertise funded by NIH R01MH110057S and the non-profit Specialists in Global Health.

### Clinical Trial

This research does not meet the definition of a clinical trial, human subjects were not prospectively assigned to one or more interventions to evaluate the effects of those interventions on health-related biomedical or behavioral outcomes. This research does involve human research conducted at the UCSF Hypoxia Research Laboratory and followed a protocol approved by UCSF Human Research Protection Program, IRB protocol 10-0043710

### Funding Statement

The work was funded by a NIH grant supplement R01MH110057-04S to SHB and a Non-Profit, Specialists in Global Health (https://sigh.global/).

### Author Declarations

UCSF Human Research Protection Program approved the research protocol. The approval number is UCSF HRPP IRB 10-00437.

